# Implementing Mandatory Testing and a Public Health Commitment to Control COVID-19 on a College Campus

**DOI:** 10.1101/2021.05.30.21258071

**Authors:** Cindy Liu, Amita Vyas, Amanda D. Castel, Karen A. McDonnell, Lynn R Goldman

## Abstract

The COVID-19 pandemic has greatly impacted US colleges and universities. As The George Washington University (GWU), a large urban university, prepared to reopen for the Fall 2020 semester, GWU established protocols to protect the health and wellness of all members of campus community. Reopening efforts included a cadre of COVID-19 surveillance systems including development of a public health COVID-19 laboratory, weekly and symptomatic SARS-CoV-2 testing and daily risk screening and symptom monitoring. Other activities included completion of a mandatory COVID-19 training and influenza vaccination for the on-campus population, quarantining of students returning to campus, campus-focused case investigations and quarantining of suspected close contacts, clinical follow-up of infected persons, and regular communication and monitoring. A smaller on-campus population of 4,435 students, faculty and staff returned to campus with later expansion of testing to accommodate GWU students living in the surrounding area. Between August 17 and December 4, 2020, 38,288 tests were performed; 220 were positive. The surveillance program demonstrated a relatively low positivity rate, with temporal clustering of infected persons mirroring community spread, and little evidence for transmission among the GWU on-campus population. These efforts demonstrate the feasibility of safely partially reopening a large urban college campus by applying core principles of public health surveillance, infectious disease epidemiology, behavioral measures, and increased testing capacity, while continuing to promote educational and research opportunities. GWU will continue to monitor the program as the pandemic evolves and periodically reassess to determine if these strategies will be successful upon a full return to in-person learning.

**Summary Box:**

1. **What is the current understanding of this subject?** When the COVID-19 pandemic began, generally universities were not prepared and it was unclear how to safely reopen colleges and universities given the uncertain risks of transmission among students, faculty and staff.
2. **What does this report add to the literature?** This case report highlights the successful approaches employed, including applying the core principles of public health surveillance and increasing testing capacity, to mitigate the spread of SARS-CoV-2 on a densely populated urban college campus resulting in a safe reopening.
3. **What are the implications for public health practice?** This experience can provide a roadmap for other universities to consider as they plan for the safe reopening of their campuses.

## INTRODUCTION

The COVID-19 pandemic reached United States (US) colleges and universities in early 2020, as students, faculty and staff found themselves in the path of the epidemic as it swept from Wuhan across regions in China and spread to Europe, especially northern Italy and Spain. ^1,2^ Universities faced many difficult decisions as students returned to campus prior to completion of their academic programs. ^3-5^ Some colleges had observed outbreaks related to spring break travel [5]. For these returning students, healthcare providers had difficulty obtaining orders for SARS-CoV-2 PCR tests which, at the time, were scarce and restricted to official US Centers for Disease Control and Prevention (CDC) purposes which did not include surveillance of asymptomatic people without known exposures.

Early reports from China described people who either did not yet manifest symptoms (presymptomatic) or were unaware they had symptoms (mildly symptomatic) or never had symptoms (asymptomatic) that were transmitting the virus yet the extent of asymptomatic transmission was unclear at the time.^6^ As COVID-19 infections and exposures were identified on campuses, the challenges to containment became clear. Students could potentially have numerous contacts on a typical college campus, whether in dorms, dining and study areas, clubs and Greek life, or in classrooms.^7-10^ Furthermore, faculty and staff, who tend to be older, could also have numerous exposures both on and off-campus and be at higher risk for severe infection.^9,11,12^ Furthermore, contact tracing was overwhelming at a time when local public health systems lacked capacity.^13^

Like most universities, The George Washington University (GWU) switched to online instruction during Spring Break (March 2020). GWU prepared to reopen the university for the Fall semester, in a manner that would protect the health and wellness of all members of the campus communities, especially those who were most vulnerable. ^14^ Physical distancing, wearing masks, shifting large classes to smaller and online formats, employee telework, de-densifying dormitory life, and establishing policies and facilities for quarantine and isolation of campus members who may have, or are confirmed to have, SARS-CoV-2 infections were among the many public health measures that were becoming standard at US colleges as they operated in the midst of a public health emergency. Despite these measures, the dense campus environment could nonetheless provide opportunities for SARS-CoV-2 transmission. ^5,8,15,16^

For these reasons GWU implemented a campus-wide public health surveillance effort for COVID-19, to include frequent SARS-CoV-2 virus testing as well as daily risk screening and symptom monitoring. No combination of interventions by universities alone can decrease viral transmission to the same extent as what may be experienced when most people have been working and studying from home. For universities to reopen, the increased risk for viral transmission must be minimized– it must be counterbalanced by the benefits to students of a college education within an on-campus environment. ^11,14^ The university had following objectives for reopening:

1. Reduce transmission within the campus environments to the extent possible.
2. Assure that the university is not accelerating transmission within the surrounding community nor placing an unreasonable burden on its healthcare systems.
3. Protect the most vulnerable, including all student, faculty, and staff with preexisting conditions.
4. Encourage telework where possible.
5. Provide online learning options for all classes so that students and faculty can isolate or quarantine when needed.

## PURPOSE

To prepare for gradual reopening of on-campus instruction, research, and administration, GWU put in place a cadre of surveillance systems for COVID-19 that took into account the challenges of: 1) understanding the transmission of the novel SARS-CoV-2, 2) setting up a university COVID-19 public health testing laboratory to conduct reliable high volume testing with a short turnaround, 3) developing and implementing a campus case investigation and campus exposure assessment team, 4) enforcing new policies and procedures to ensure the health and safety of the university and surrounding community, 5) and communicating and disseminating results to the public. This paper describes the approach that was taken.

## METHODS

### COVID-19 Surveillance Methods

The first step in GW’s development of a reopening plan was to develop the methods to be employed for COVID-19 surveillance. Public health surveillance of SARS-CoV-2, was done to:

1. *Track* the *prevalence* of the disease over time so that action can be taken to intervene if and when there is a resurgence.
2. *Isolate cases* of COVID-19 to prevent spread.
3. *Quarantine* all close *contacts* of COVID-19 cases, test for the SARS-CoV-2 and prevent further spread.
4. *Monitor risks such as travel and exposures to non-campus venues and persons to prevent campus spread*.

**Table 1** summarizes approaches for public health surveillance of COVID-19 and GW’s approach to them.

**Table 1.** COVID-19 Public Health Surveillance and Corresponding GWU Approach.

| Public Health Considerations | Evidence-base/Standard Definition | GWU approach |
| --- | --- | --- |
| **Case definitions<sup>26</sup>** | • **Clinical**--- symptom clusters plus no alternative diagnosis; or<br>• **Laboratory**— detection of SARS-Cov-2 RNA or antigen in clinical specimens<br>• **Epidemiologic**- linkage to place, person or circumstance with known COVID-19 risk<br>• **Vital Records**-cause of death | • **Clinical** -- symptoms consistent with COVID-19 minus an alternative diagnosis as determined by Student/Occupational Health (Probable)<br>• **Laboratory** --positive PCR result from GWU Public Health Lab (Confirmed)<br>• **Epidemiologic** - linkage to confirmed campus case based on travel, close contact or membership in a campus cohort among a person with symptoms of COVID-19 (Epidemiologic) |
| **Symptom self-monitoring** | • Mild and moderate COVID-19 symptoms are quite similar to the symptoms of a myriad of other acute viral illnesses.<br>• No unique pattern of symptoms distinguishes COVID-19 from many other acute illnesses.<br>• Symptom-based criteria must be accompanied by a clinical determination of “no alternative more likely diagnosis”<br>• Medical encounter is required in order to identify a case on the basis of symptom self-monitoring. | • Instituted daily symptom self-monitoring through PointnClick electronic health record.<br>• Daily 5-question survey automatically generated for on-campus population<br>• Persons reporting any symptom of COVID-19 referred to Student or Occupational health for additional screening and telehealth visit if needed |
| **Temperature checks** | • Frequently used to diagnose COVID-19<br>• Usually performed with a handheld IR (infrared) device or a camera IR device, sometimes in concert with a symptom questionnaire. | • Based on lack of evidence, did not institute routine temperature checks for on-campus population |
|  | • Systematic review found method is “unfavorable” Handheld IR thermometers have measurement inaccuracies, low sensitivity and are prone to operator error <sup>17</sup> |  |
| **Virus tests** | • Viral nucleic acid or viral protein tests can support the diagnosis of COVID-19 via direct detection of the virus in nasopharyngeal, nasal or salivary secretions.<br>• Currently no FDA-approved tests for COVID; however, but testing may be performed under FDA Emergency Use Authorizations (EUAs). <sup>18</sup> | • Developed PCR based viral assay<br>• EUA obtained prior to start of return to campus<br>• Anterior nasal specimens collected by provider<br>• Tests processed in newly formed Public Health Laboratory<br>• Results provided in 8-20 hours |
| **Antibody diagnostic tests** | • Presence of antibodies to SARS-CoV-19 indicates history of exposure to the virus. >99% of those with a positive PCR test will become seropositive by 14 days after symptom onset, but the persistence of anti-SARS-CoV-19 antibodies is still little understood. | • Use of IgG/IgM antibody testing for persons who test positive but report a history of prior infection. |

### GWU Approach to on-Campus COVID-19 Surveillance

Based on the epidemiology as it was evolving, including early signs that college campus environments could have relatively high COVID-19 transmission, a combination of approaches was taken:

- Agreement to participate in campus COVID-related activities including completion of a mandatory training about COVID-19 and adherence to requirements to wear face coverings, practice physical distancing, and to not hold gatherings of >10 people.
- Mandatory periodic (*weekly*) COVID-19 virus testing as well as daily symptom monitoring for all on-campus students, faculty, and staff.
- Return to campus monitoring and testing for on-campus residential students on arrival and five days later. Quarantine of residence hall students pending two negative PCR tests.
- On-campus investigations to identify transmission early and quarantine suspected close contacts of cases.
- Clinical follow-up, quarantine and testing at any point for any member of the on-campus community who develops symptoms.
- Per rules of the District of Columbia Health Department a 14-day quarantine of anyone returning to campus from states defined by them as “high transmission”.
- Flu vaccine at a university clinic, or documentation of a flu shot obtained elsewhere. Of these, all were carried out all successfully except the pre-campus testing.

### Development of the GWU SARS-CoV-2 PCR Test

This is the centerpiece of this effort. One of us (C.L.) obtained IRB approval for a research protocol to initiate COVID-19 studies. In the process of conducting this research, the laboratory experimented with a number of approaches to develop a PCR test kit and workflow that met a number of *a priori* requirements: (1) use of a PCR test, the gold standard for sensitivity; (2) an anterior nasal swab sampling procedure which is acceptable to the GWU community; (3) automation to support high test volumes and rapid test turnaround time (<24 hours); and (4) reliance on off-the-shelf reagents and consumables to minimize supply chain problems.^16,19^

After setting up a CLIA high-complexity testing laboratory, the GWU Emergency Use Authorization (EUA) for the GWU COVID-19 PCR test was approved 7/29/2020. The test has high positive agreement (95%) and negative agreement (100%) with other EUA tests, a must for higher scale screening of people with no or minimal symptoms. Given that the original EUA required provider-administered tests, health providers were needed from GWU’s medical faculty and nursing school, as well as PPE, to collect samples. At the start of the fall semester, additional studies were underway to collect data for additional claims, such as self-collection, pooling samples, and asymptomatic testing.

### Test Ordering and Return of Results

The objective was to establish a data system that would seamlessly flow the PCR results to individuals, clinicians and public health agencies, and integrate this information with symptom monitoring data. The short preparation time and relative unavailability of vendors in Summer 2020 led us to develop a basic yet highly customized laboratory information management system and to utilize a COVID-19 module under development from an existing electronic health record system, PointNClick® (PnC), that was already deployed in the student health service (the Colonial Health Center). Testing appointments are made on PnC and each appointment is linked to a unique QR code. At test check-in, scanning the appointment QR code and the barcode on pre-printed specimen tubes links the specimen to the test order. Test results are reviewed by the Lab Director and results are released to PnC for clinician acknowledgement. Once acknowledged, members of the community are notified by email that they can review their test results by logging into PnC. PnC also facilitates reporting results, along with any other required data to state and local health departments.

### Case Investigations and Campus Exposure Assessment

GWU epidemiological and behavioral science faculty led the development and implementation of the Campus COVID Support Team (CCST) to manage and enforce risk screening and symptom monitoring; case investigation; case reporting to the local health departments; on-campus contact identification and exposure assessment; rapid isolation of cases and/or quarantine of contacts; and care for the worried well as well as those in quarantine or isolation. The CCST opens a case investigation for each person with a positive test; an initial contact within a few hours puts into motion a sequence of steps that includes review of the isolation protocol, a schedule of telehealth consultations, and a review of potential campus exposures (spaces and persons). The CCST coordinates with teams across the university to ensure that spaces are cleaned according to CDC COVID-19 cleaning guidelines^11^; students have academic support as well as food and other necessities; employees have documentation for pandemic leave; and access to buildings and spaces are restricted for isolation and quarantine periods.

### Communication and Monitoring

As required lab reports are reported electronically to DC and VA public health departments. Summary report are given several times a week to university leadership. Special communications provide information to those who are participating in the on-campus testing both about the results but also to remind them about their responsibilities in complying with on-campus requirements. A public facing website describes the test protocols and a COVID-19 Dashboard (https://coronavirus.gwu.edu/dashboard) provides daily updates on numbers of tests, test-positives and test positivity rates, by campus and for students versus faculty and staff.

Additionally, the campus closely monitors situational surveillance and epidemiologic metrics. These include numbers of persons in quarantine, the campus-specific 7-day average positivity, the median numbers of contacts per case, and the size and detection of potential outbreaks. These metrics, in combination with those presented in **Figure 1** are used to monitor the campus status and the need to adjust layers of prevention and mitigation as necessary.

**Figure 1:**
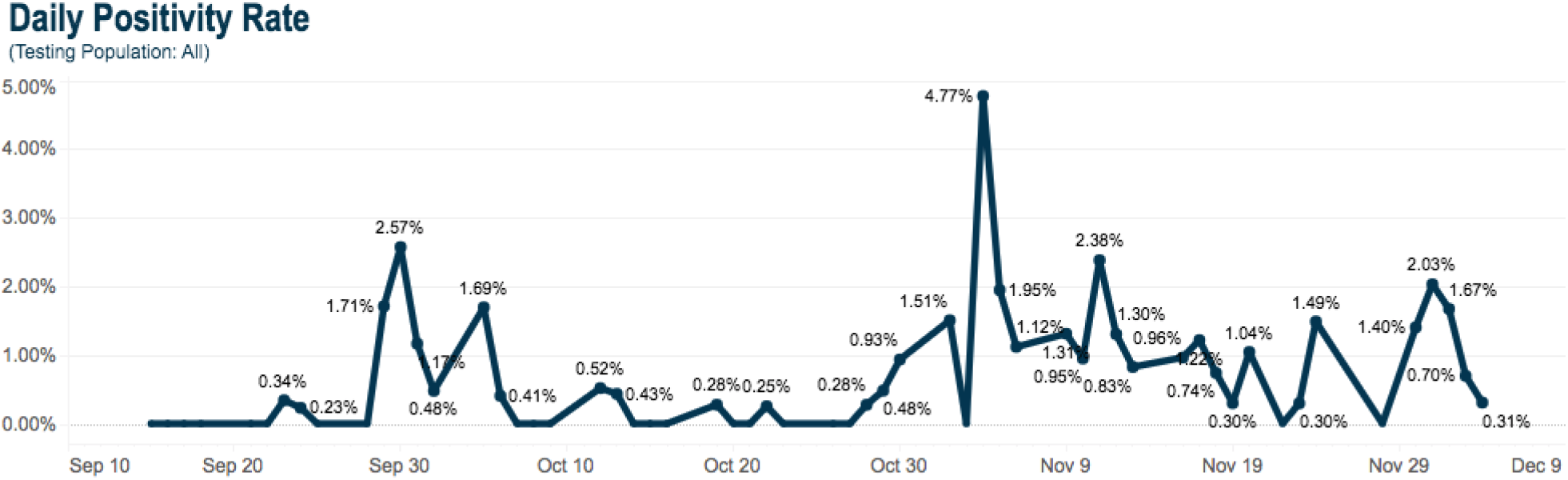
Daily Positivity Rate August 17-December 4, 2020.

### Consent

The GWU Institutional Review Board determined that this project is not human subjects research because this is a surveillance project and not a systematic investigation designed to contribute to generalizable knowledge. Nonetheless, all participants were asked to sign a consent form giving permission for viral testing, the disclosure of viral test results to the District of Columbia Department of Health as well as to student or employee health services, and the GWU Campus COVID Support Team (CCST).

## OUTCOMES

### GWU Early Results of on-Campus COVID-19 Surveillance

A total of 1,308 students (∼500 residing on campus), 1,643 staff, 534 faculty and ∼500 contractors were included in the return to campus cohort. This resulted in an on-campus population of 4,435 to be tested weekly (considerably less than the 25,000 originally anticipated.) However, as the pandemic progressed, the test site expanded to accommodate all GWU students living in the surrounding metropolitan area. For context, of about 28,000 students enrolled in Fall (including in online programs), an estimated 15,000 were in DC, MD or VA with about 6,280 residing in DC. Among these, around 440 were living in the Foggy Bottom/ west End neighborhood immediately surrounding GWU (and not in the residence halls).

**Figure 1** shows the numbers of positive tests identified by day through the Fall Semester, from when testing began August 17, 2020 and through December 4. During this period there were a total of 38,288 tests performed (21,573 for students and 16,715 for employees) of which 220 were positive (175 students for a positivity rate of 0.81% (175/21573) and 45 for employees for a positivity rate of 0.27% (45/16715)). Temporal clusters of positive cases mirrored community spread with increases in cases for holiday gatherings, e.g., the spike in test positivity that occurred shortly after Halloween. However, serial surveillance demonstrated a significantly lower positivity rate on campus than national statistics and little evidence for transmission among the on-campus population.

## LESSONS LEARNED

In this mid-pandemic era, GWU has been able to safely and effectively reopen its campus to ensure the safety of its community while continuing to promote educational and research opportunities. The development of rapid and high-throughput PCR-based COVID-19 testing is feasible in a CLIA laboratory setting with mostly off-the shelf reagents and consumables. Such surveillance requires adequate testing capacity as well as a rapid turnaround time (<24 hours) to identify persons testing positive for SARS-CoV-2 and thereby mitigate the spread of the virus throughout the campus population. As GWU gradually opens the campus to additional members of the GWU community, ensuring the ability to maintain lab capacity and results notification on a timely basis will remain essential, including the possibility of using pooled testing.^20^

This overall approach to developing a public health surveillance program in a large urban college campus is recommended especially in the context of global universities that are densely populated and have on-campus student housing. This type of surveillance should be viewed as a backstop to all other efforts to provide safe campus environments and create new norms for health protective behaviors and in no way replaces those efforts. By monitoring compliance amongst the on-campus population it will be possible to determine if the cadence for surveillance testing should be modified based on positivity rates among specific subpopulations, and measure behavioral compliance for social distancing fatigue and wearing of face coverings.

It is uncertain whether this strategy would have successful through the year if the campus had been fully populated. Some models and guidelines have suggested that biweekly monitoring may be necessary to halt transmission of SARS-CoV-2. ^21,22^ This approach would allow GWU to transition to twice weekly testing if an uptick of transmission occurs on the campus and/or in the surrounding community. Another unknown is that there is very little information on how testing and other efforts to slow transmission impact behavioral prevention efforts. More data are needed to understand the extent behavior is modified under the circumstances of mandatory testing, symptom tracking and requirements for masks and distancing.^23-25^

In conclusion, the approach taken to safely reopen the GWU campus applied core principles of public health surveillance, infectious disease epidemiology and transmission, behavioral measures, and increased testing capacity. GWU met the goals of reducing transmission on campus without further burdening already strained public health and healthcare resources in the wider community. Further, these efforts demonstrated the feasibility of implementing on-campus surveillance and public health control of COVID-19 utilizing high throughput testing to rapidly obtain test results, analyze these data in real-time, and to make informed decisions on behalf of individuals as well as campus communities. In Spring and beyond, efforts include expanding these efforts in other settings, the integration of mass vaccination clinics into the workflow, and continuing to evaluate and monitor the impact these efforts.

## Data Availability

These data are not publicly available. Aggregate testing data can be accessed on the GWU COVID-19 Testing Dashboard available at https://coronavirus.gwu.edu/dashboard

## Acknowledgements

We would like to thank members of The George Washington University COVID-19 Public Health Laboratory (Maliha Aziz, Melody Fung, Daniel Park, Vanessa Quinlivan, Juan Salazar, David Villani), Colonial Health Center (Dr. Isabel Goldenberg), Occupational Health (Dr. Raymond Lucas), Office of General Counsel (Charles Barber), Campus Safety and Security (Scott Burnotes and Kate Fox) and Campus COVID Support Team (Nitasha Nagaraj, Patrick Beane, Nigussie Gemechu, Gary Sardon, Emily Weiss, Mira Agneshwar, Jasmine Jones, Cassidy Spence) for their support and assistance with COVID-19 reopening and response efforts.

## Notes

### Competing Interest Statement

The authors have declared no competing interest.

### Funding Statement

This work was funded by the George Washington University. No external funding was received.

### Author Declarations

The George Washington University Institutional Review Board reviewed and determined this to be non-human subjects research. This determination was made because this is a surveillance project and is not a systematic investigation designed to contribute to generalizable knowledge.

